# Use of SGLT2i and ARNi in patients with atrial fibrillation and heart failure in 2021-2022: an analysis of real-world data

**DOI:** 10.1101/2023.09.08.23295280

**Authors:** Alvaro Alonso, Alanna A. Morris, Ashley I. Naimi, Aniqa B. Alam, Linzi Li, Vinita Subramanya, Lin Yee Chen, Pamela L. Lutsey

## Abstract

**Objective:** To evaluate utilization of sodium-glucose cotransporter-2 inhibitors (SGLT2i) and angiotensin receptor neprilysin inhibitors (ARNi) in patients with atrial fibrillation (AF) and heart failure (HF).

**Methods:** We analyzed the MarketScan databases for the period 1/1/2021 to 6/30/2022. Validated algorithms were used to identify patients with AF and HF, and to classify patients into HF with reduced ejection fraction (HFrEF) or preserved ejection fraction (HFpEF). We assessed the prevalence of SGLT2i and ARNi use overall and by HF type. Additionally, we explored correlates of lower utilization, including demographics and comorbidities.

**Results:** The study population included 60,927 patients (mean age 75, 43% female) diagnosed with AF and HF (85% with HFpEF, 15% with HFrEF). Prevalence of ARNi use was 11% overall (30% in HFrEF, 8% in HFpEF), while the corresponding figure was 6% for SGLT2i (13% in HFrEF, 5% in HFpEF). Use of both medications increased over the study period: ARNi from 9% to 12% (from 22% to 29% in HFrEF, from 6% to 8% in HFpEF), and SGLT2i from 3% to 9% (from 6% to 16% in HFrEF, from 2% to 7% in HFpEF). Female sex, older age, and specific comorbidities were associated with lower utilization of these two medication types overall and by HF type.

**Conclusion:** Use of ARNi and SGLT2i in patients with AF and HF is suboptimal, particularly among females and older individuals, though utilization is increasing. These results underscore the need for understanding reasons for these disparities and developing interventions to improve adoption of evidence-based therapies among patients with comorbid AF and HF.

## BACKGROUND

Atrial fibrillation (AF) and heart failure (HF) are two commonly occurring cardiovascular conditions that frequently coexist and interact with each other.^1^ More than 30% of AF patients present with HF,^2^ and >25% of HF patients with New York Heart Association functional class III-IV also have AF.^3^ Moreover, the coexistence of HF and AF often worsen patient’s symptoms and disease progression, leading to increased morbidity and mortality.^4^ Comorbid AF and HF can also contribute to challenges in clinical management given the high burden of multimorbidity and polypharmacy in these patients.^5-8^ Despite the large burden of cooccurring HF and AF, treatment guidelines offer insufficient guidance about best approaches to manage this group of patients.^9, 10^

Based on results from landmark randomized trials testing the efficacy of sodium-glucose co-transporter 2 inhibitors (SGLT2i) and angiotensin receptor-neprilysin inhibitors (ARNi) in the treatment of HF, current treatment guidelines strongly recommend the use of SGLT2i and ARNi in the management of patients with HF with reduced ejection fraction (HFrEF), HF with mildly reduced ejection fraction (HFmrEF), and HF with preserved ejection fraction (HFpEF).^9, 11^ Secondary analyses of large SGLT2i and ARNi trials showed no difference in efficacy between patients with AF and those without AF.^12, 13^ However, these trials were not powered to detect such differences. Also, it is unclear whether findings accurately generalize to real-world patient populations given the restrictive inclusion criteria of many clinical trials. These knowledge gaps could potentially hinder the appropriate use of SGLT2i and ARNi in patients with comorbid AF and HF.^14^ To gain a better understanding of the utilization of SGLT2i and ARNi in this patient group, we examined the frequency of prescription and the demographic and clinical factors associated with SGLT2i and ARNi use in a large contemporary cohort of patients with comorbid AF and HF identified from a healthcare claims database in the United States.

## METHODS

### Data sources: MarketScan databases

We identified patients with AF and HF included in the Merative MarketScan Commercial and Medicare databases (Merative, Ann Arbor, MI). The MarketScan databases include patient-level information on clinical utilization in inpatient and outpatient settings, as well as enrollment and prescription data, from individuals enrolled in a selection of large employers, health governments, and government and public health organizations in the US. The MarketScan Commercial database contains data from individuals insured through employer-sponsored plans, while the MarketScan Medicare database includes Medicare-eligible individuals with Medicare Supplemental or Medicare Advantage plans. For the current study, we considered the period January 1, 2021 to June 30, 2022 to evaluate contemporary utilization after approval by the US Food and Drug Administration in 2020 of SGLT2i for HFrEF treatment in patients with and without type 2 diabetes (approval of SGLT2i for HFpEF treatment occurred in early 2022; off-label use may have occurred prior), and the approval of the ARNi sacubitril/valsartan to patients with chronic HF independently of their EF in 2021.

The Institutional Review Board (IRB) of Emory University determined that this study did not require IRB review because it does not meet the definition of research with “human subjects” or “clinical investigation” because the database provides researchers with deidentified information.

### Identification of patients with comorbid AF and HF

Ascertainment of AF was done following previously described algorithms.^15^ Briefly, AF diagnosis required the presence of ICD-10-CM code I48.xx in two outpatient claims more than 7 days and less than 365 days apart or in one inpatient claim. Diagnosis of HF required one inpatient claim with HF as primary discharge diagnoses or two inpatient claims with HF diagnoses in any position or two outpatient claims with HF diagnoses in any position more than 7 days and less than 365 days apart (ICD-10-CM codes I09.81, I11.0, I13.0, I13.2, I50.xx).^16^ We further categorized HF cases at the time of first diagnosis as HFrEF or HFpEF using a validated algorithm.^17, 18^ This algorithm considers demographic variables and inpatient, outpatient and pharmacy claims in the period 6 months prior and 1 month after the diagnosis of HF, and in the MarketScan databases has a positive predictive value of 73% and 81% for HFrEF and HFpEF, respectively.^18^ Date of comorbid AF/HF diagnosis was defined as the latest of the diagnosis of AF or the diagnosis of HF.

### Medication use

The primary endpoint variable was the use of SGLT2i and ARNi. Filled outpatient prescriptions for sacubitril/valsartan (ARNi) and dapagliflozin and empaglifozin (SGLT2i approved for HF treatment) during the study period were identified. Information on the following medications was also ascertained: beta-blockers, mineralocorticoid receptor antagonists (MRAs), angiotensin converting enzyme inhibitors (ACEI), angiotensin receptor blockers (ARB), anti-arrhythmic agents, and oral anticoagulants. We determined use of a specific medication at the time of AF/HF diagnosis if a filled prescription for a specific medication was active 90 days before or after the diagnosis date. We defined triple therapy if a patient was receiving a beta-blocker, an MRA, and an ACEi, ARB or ARNi, and quadruple therapy if the patient also received a SGLT2i.

### Comorbidities

Presence of comorbidities at the time of AF/HF diagnosis was determined from International Classification of Disease, 10^th^ edition, Clinical Modification (ICD-10-CM) diagnosis codes in outpatient and inpatient claims. Conditions were defined using algorithms proposed by the Chronic Conditions Data Warehouse from the Center for Medicare and Medicaid Services (https://www2.ccwdata.org/web/guest/condition-categories-chronic). These comorbidities were selected based on being potential predictors of ARNi or SGLT2i use.

### Statistical analysis

We evaluated patient characteristics at time of AF/HF diagnosis, overall and by HF type (HFrEF and HFpEF). Quarterly prevalence of use of ARNi and SGLT2i over the study period was calculated as the proportion of patients with prevalent AF/HF in that quarter who had at least one active prescription for that medication during that time period. We calculated proportions of patients using ARNi and SGLT2i around the time of AF/HF diagnosis across demographic and clinical variables. Finally, we evaluated independent demographic and clinical predictors of ARNi and SGLT2i use by calculating relative risks from log-binomial models (or Poisson models with robust variance estimation if the log-binomial model did not converge). These analyses included the following covariates simultaneously in the models: age, sex, HF type, hypertension, diabetes, hyperlipidemia, coronary artery disease, ischemic stroke, hemorrhagic stroke, chronic kidney disease, chronic obstructive pulmonary disease, use of oral anticoagulants, use of antiarrhythmic drugs, ACEi/ARB, beta-blocker, MRA, and, when corresponding, ARNi and SGLT2i. These variables were selected because they may directly influence use of ARNi and SGLT2i, or be correlates of other independent factors affecting ARNi or SGLT2i use. Analyses were conducted in the overall AF/HF population, by HF type and, for SGLT2i, by diabetes status.

## RESULTS

We identified 60,927 patients diagnosed with comorbid AF and HF enrolled in the MarketScan databases during the period January 1, 2021 and June 30, 2022. Of these, 9,335 (15%) were categorized as HFrEF and 51,592 (85%) as HFpEF. Patient characteristics overall and by HF type are presented in Table 1. Mean age was 75 years in the overall sample, and 68 and 76 among patients with HFrEF and HFpEF, respectively. Over three quarters of patients with HFrEF were male, while numbers of males and females was similar in HFpEF. Prevalence of comorbidities was high overall and in both HF types.

**Table 1.** Characteristics of patients with atrial fibrillation and heart failure at the time of co-diagnosis, MarketScan 2021-2022.

|  | Overall | HFrEF | HFpEF |
| --- | --- | --- | --- |
| N | 60,927 | 9,335 | 51,592 |
| Age, years | 75 (13) | 68 (14) | 76 (12) |
| Sex |  |  |  |
| Female | 26,291 (43%) | 2,281 (24%) | 24,010 (47%) |
| Male | 34,636 (57%) | 7,054 (76%) | 27,582 (53%) |
| Hypertension | 57,122 (94%) | 8,310 (89%) | 48,812 (95%) |
| Diabetes | 27,573 (45%) | 3,811 (41%) | 23,762 (46%) |
| Hyperlipidemia | 46,580 (76%) | 6,704 (72%) | 39,876 (77%) |
| Coronary artery disease | 37,308 (61%) | 6,129 (66%) | 31,179 (60%) |
| Chronic kidney disease | 28,550 (47%) | 3,833 (41%) | 24,717 (48%) |
| Ischemic stroke | 6,041 (10%) | 763 (8%) | 5,278 (10%) |
| Intracranial bleeding | 1,072 (2%) | 136 (1%) | 936 (2%) |
| COPD | 19,121 (31%) | 2,192 (23%) | 16,929 (33%) |
| Oral anticoagulant use | 45,262 (74%) | 7,184 (77%) | 38,078 (74%) |
| Antiarrhythmic drug use | 13,899 (23%) | 3,145 (34%) | 10,754 (21%) |

Values correspond to N (%) or mean (standard deviation). COPD: chronic obstructive pulmonary disease. HFpEF: heart failure with preserved ejection fraction. HFrEF: heart failure with reduced ejection fraction.

Prevalence of ARNi use around the time of AF/HF diagnosis was 11%, 30% in HFrEF and 8% in HFpEF. Corresponding figures for SGLT2i were 6% (overall), 13% (HFrEF) and 5% (HFpEF). Only 31% of HFrEF patients were receiving triple therapy (beta blocker plus ACEi/ARB/ARNi plus MRA). Prevalence of quadruple therapy was 7% in HFrEF (Table 2). SGLT2i use was higher in patients with diabetes (11%) than in those without diabetes (3%) (Supplemental Figure 1).

**Table 2.** Prevalence of guideline-directed medical therapy active prescriptions in patients with comorbid AF and HF at the time of co-diagnosis (+/-90 days), MarketScan 2021-2022.

|  | Overall (N = 60,927) | HFrEF (N = 9,335) | HFpEF (N = 51,592) |
| --- | --- | --- | --- |
| Beta blocker | 48,330 (79%) | 8,312 (89%) | 40,018 (78%) |
| ACEi/ARB | 30,013 (49%) | 5,055 (54%) | 24,958 (48%) |
| ARNi | 6,856 (11%) | 2,773 (30%) | 4,083 (8%) |
| ACEi/ARB/ARNi | 34,896 (57%) | 6,948 (74%) | 27,948 (54%) |
| MRA | 13,167 (22%) | 3,505 (38%) | 9,662 (19%) |
| SGLT2i | 3,828 (6%) | 1,178 (13%) | 2,650 (5%) |
| Triple therapy | 8,279 (14%) | 2,851 (31%) | 5,428 (11%) |
| Quadruple therapy | 1,446 (2%) | 663 (7%) | 783 (2%) |

ACEi: angiotensin converting enzyme inhibitor. ARB: angiotensin receptor blocker. ARNi: angiotensin receptor-neprilysin inhibitor. HFpEF: heart failure with preserved ejection fraction. HFrEF: heart failure with reduced ejection fraction. MRA: mineralocorticoid receptor antagonist. SGLT2i: sodium-glucose cotransporter-2 inhibitor. Triple therapy: use of ACEi, ARB or ARNi plus beta blocker plus MRA.
Quadruple therapy: triple therapy plus use of SGLT2i.

During the study period, use of ARNi slightly increased from 9% in the first quarter of 2021 to 12% in the second quarter of 2022 (Figure 1). Trends showing increased use were similar for HFrEF and HFpEF, with increases from 22% to 29% in HFrEF and from 6% to 8% in HFpEF. In contrast, use of SGLT2i experienced a large relative increase, with prevalence almost tripling, from 3% to 9% for all HF, and from 6% to 16% in HFrEF and 2% to 7% in HFpEF (Figure 1, Supplemental Table 1).

**Figure 1.**
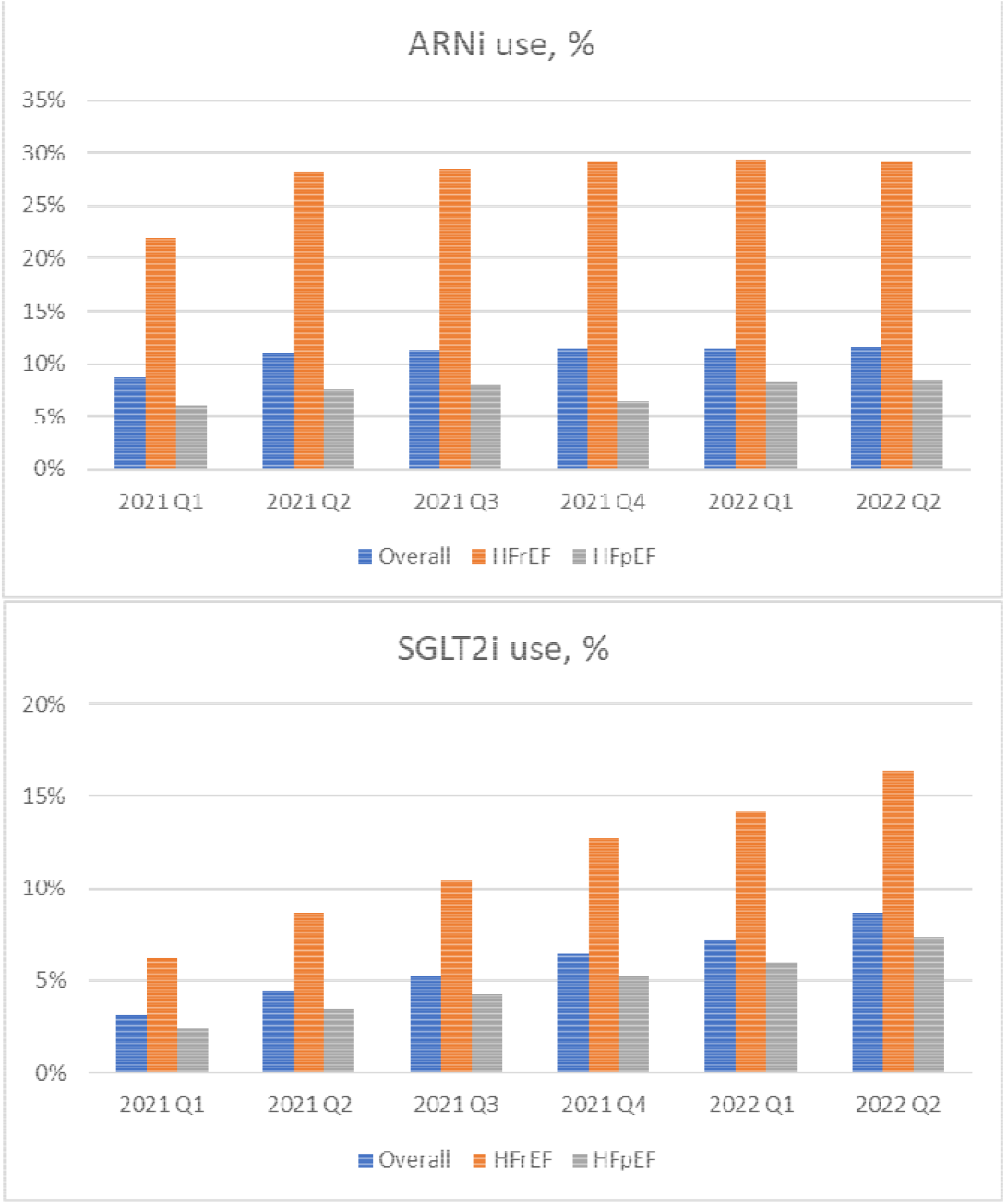
Prevalence of sodium-glucose cotransporter-2 inhibitors and angiotensin receptor-neprilysin inhibitor use among patients with atrial fibrillation and heart failure by quarter, MarketScan databases 2021-2022. ARNi: angiotensin receptor-neprilysin inhibitor; HFpEF: heart failure with preserved ejection fraction; HFrEF: heart failure with reduced ejection fraction; SGLT2i: sodium-glucose cotransporter-2 inhibitor.

Figure 2 shows the use of ARNi and SGLT2i by age, sex, and HF type. Female and older patients had lower prevalence of use of both medication types. Similarly, we calculated prevalence of use of ARNi and SGLT2i by presence of comorbidities and use of selected medications, finding lower use of ARNi and SGLT2i in patients with some comorbidities and higher use in those receiving other medications for AF or HF (Figure 3). The pattern was similar by HF type (HFrEF and HFpEF), with overall lower frequency of use in patients with HFpEF than in those with HFrEF (Supplemental Figure 2). In multivariable analyses, several demographic and clinical factors were strong predictors of ARNi and SGLT2i use in the overall HF population (Table 3). Older age and female sex were associated with lower probability of being prescribed these medications. Compared to patients younger than 65, patients 85 and older had a 58% and 76% lower probability of being prescribed ARNi or SGLT2i, respectively. Quadruple therapy was 91% less likely in 85 and older patients than those younger than 65. Female patients were 32% and 27% less likely than male counterparts to receive ARNi or SGLT2i, respectively, than their male counterparts. Patients with prior history of some comorbidities, including diabetes, ischemic and hemorrhagic stroke, chronic kidney disease and chronic obstructive pulmonary disease, were less likely to be prescribed ARNi. Hypertension, ischemic and hemorrhagic stroke, and chronic obstructive pulmonary disease were also associated with lower likelihood of SGLT2i prescription. Patterns were similar for prescriptions of ARNi and SGLT2i in HFrEF and HFpEF separately (Supplemental Table 2) and for prescriptions of SGLT2i by diabetes status (Supplemental Table 3).

**Table 3.**
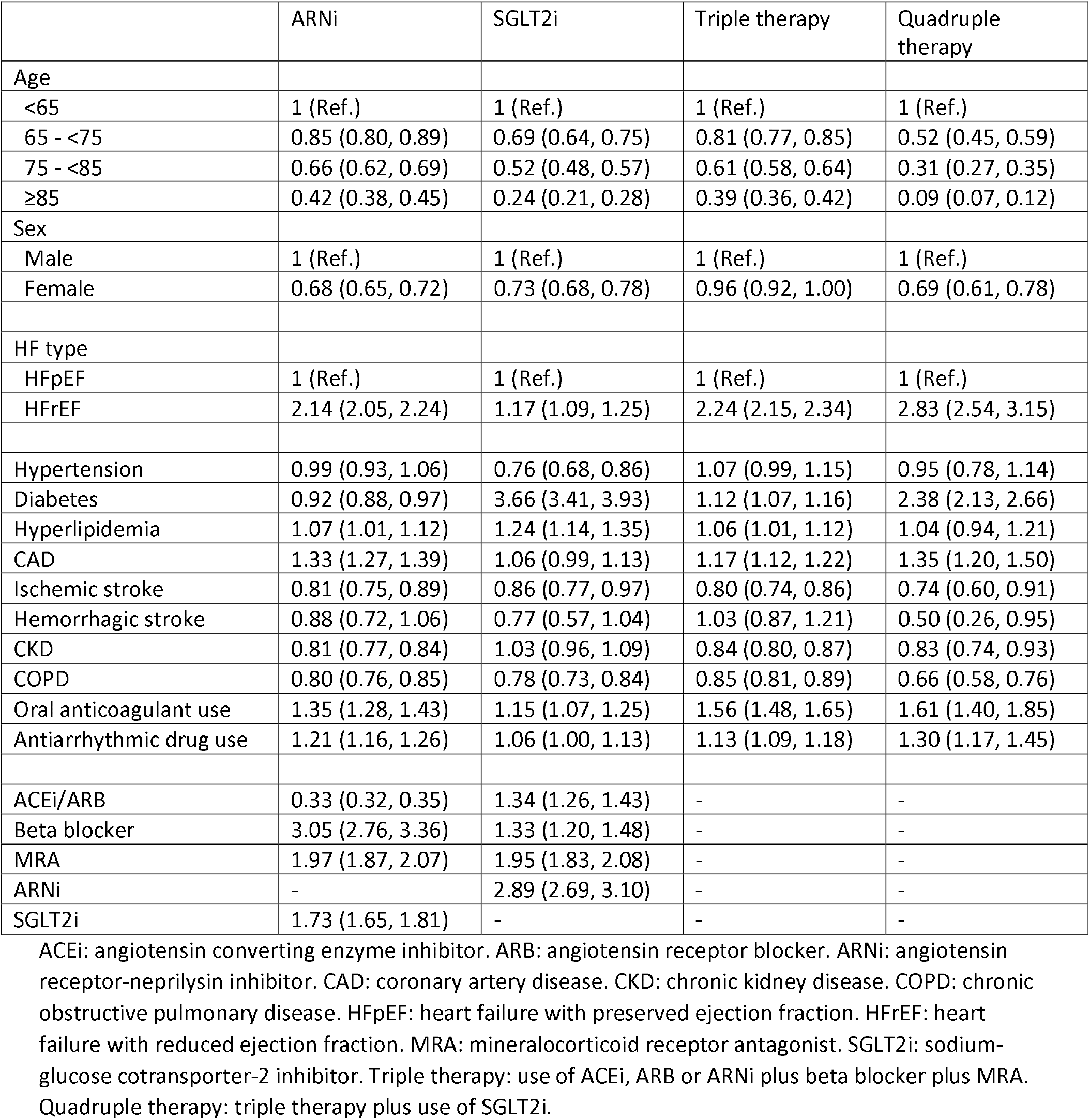
Predictors of use of angiotensin receptor-neprilysin inhibitors, sodium-glucose cotransporter-2 inhibitors, triple therapy and quadruple therapy in patients with AF and HF at time of co-diagnosis (+/-90 days). Results correspond to relative risks and 95% confidence intervals from log-linear model (or Poisson regression when model not converging) including all variables in the table. MarketScan, 2021-2022.

|  | ARNi | SGLT2i | Triple therapy | Quadruple therapy |
| --- | --- | --- | --- | --- |
| Age |  |  |  |  |
| <65 | 1 (Ref.) | 1 (Ref.) | 1 (Ref.) | 1 (Ref.) |
| 65 - <75 | 0.85 (0.80, 0.89) | 0.69 (0.64, 0.75) | 0.81 (0.77, 0.85) | 0.52 (0.45, 0.59) |
| 75 - <85 | 0.66 (0.62, 0.69) | 0.52 (0.48, 0.57) | 0.61 (0.58, 0.64) | 0.31 (0.27, 0.35) |
| ≥85 | 0.42 (0.38, 0.45) | 0.24 (0.21, 0.28) | 0.39 (0.36, 0.42) | 0.09 (0.07, 0.12) |
| Sex |  |  |  |  |
| Male | 1 (Ref.) | 1 (Ref.) | 1 (Ref.) | 1 (Ref.) |
| Female | 0.68 (0.65, 0.72) | 0.73 (0.68, 0.78) | 0.96 (0.92, 1.00) | 0.69 (0.61, 0.78) |
| HF type |  |  |  |  |
| HFpEF | 1 (Ref.) | 1 (Ref.) | 1 (Ref.) | 1 (Ref.) |
| HFrEF | 2.14 (2.05, 2.24) | 1.17 (1.09, 1.25) | 2.24 (2.15, 2.34) | 2.83 (2.54, 3.15) |
| Hypertension | 0.99 (0.93, 1.06) | 0.76 (0.68, 0.86) | 1.07 (0.99, 1.15) | 0.95 (0.78, 1.14) |
| Diabetes | 0.92 (0.88, 0.97) | 3.66 (3.41, 3.93) | 1.12 (1.07, 1.16) | 2.38 (2.13, 2.66) |
| Hyperlipidemia | 1.07 (1.01, 1.12) | 1.24 (1.14, 1.35) | 1.06 (1.01, 1.12) | 1.04 (0.94, 1.21) |
| CAD | 1.33 (1.27, 1.39) | 1.06 (0.99, 1.13) | 1.17 (1.12, 1.22) | 1.35 (1.20, 1.50) |
| Ischemic stroke | 0.81 (0.75, 0.89) | 0.86 (0.77, 0.97) | 0.80 (0.74, 0.86) | 0.74 (0.60, 0.91) |
| Hemorrhagic stroke | 0.88 (0.72, 1.06) | 0.77 (0.57, 1.04) | 1.03 (0.87, 1.21) | 0.50 (0.26, 0.95) |
| CKD | 0.81 (0.77, 0.84) | 1.03 (0.96, 1.09) | 0.84 (0.80, 0.87) | 0.83 (0.74, 0.93) |
| COPD | 0.80 (0.76, 0.85) | 0.78 (0.73, 0.84) | 0.85 (0.81, 0.89) | 0.66 (0.58, 0.76) |
| Oral anticoagulant use | 1.35 (1.28, 1.43) | 1.15 (1.07, 1.25) | 1.56 (1.48, 1.65) | 1.61 (1.40, 1.85) |
| Antiarrhythmic drug use | 1.21 (1.16, 1.26) | 1.06 (1.00, 1.13) | 1.13 (1.09, 1.18) | 1.30 (1.17, 1.45) |
| ACEi/ARB | 0.33 (0.32, 0.35) | 1.34 (1.26, 1.43) | - | - |
| Beta blocker | 3.05 (2.76, 3.36) | 1.33 (1.20, 1.48) | - | - |
| MRA | 1.97 (1.87, 2.07) | 1.95 (1.83, 2.08) | - | - |
| ARNi | - | 2.89 (2.69, 3.10) | - | - |
| SGLT2i | 1.73 (1.65, 1.81) | - | - | - |

ACEi: angiotensin converting enzyme inhibitor. ARB: angiotensin receptor blocker. ARNi: angiotensin receptor-neprilysin inhibitor. CAD: coronary artery disease. CKD: chronic kidney disease. COPD: chronic obstructive pulmonary disease. HFpEF: heart failure with preserved ejection fraction. HFrEF: heart failure with reduced ejection fraction. MRA: mineralocorticoid receptor antagonist. SGLT2i: sodium-glucose cotransporter-2 inhibitor. Triple therapy: use of ACEi, ARB or ARNi plus beta blocker plus MRA. Quadruple therapy: triple therapy plus use of SGLT2i.

**Figure 2.**
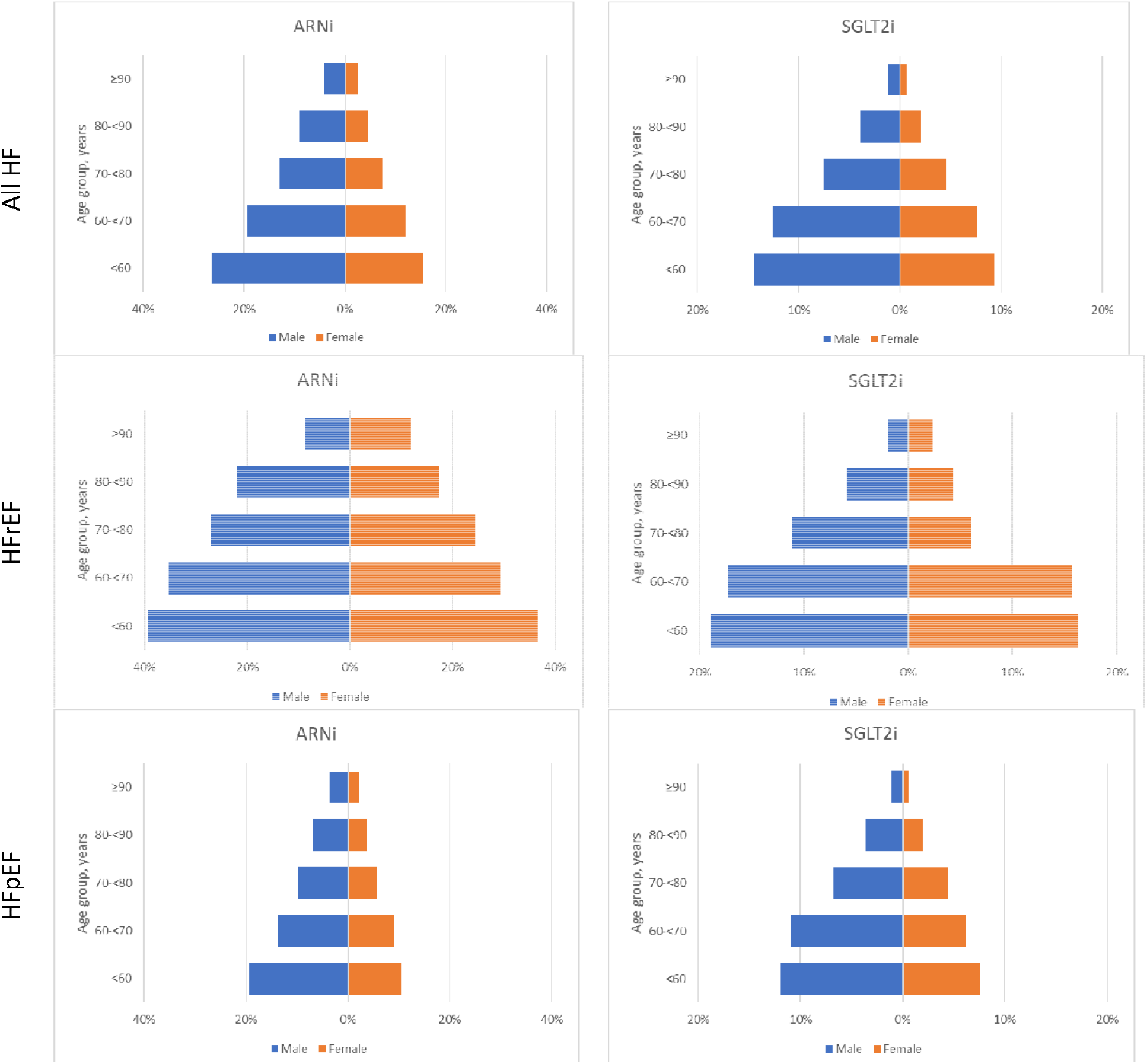
Prevalence of angiotensin receptor-neprilysin inhibitor and sodium-glucose cotransporter-2 inhibitor use among patients with atrial fibrillation and heart failure by age and sex, MarketScan databases 2021-2022. ARNi: angiotensin receptor-neprilysin inhibitor; HFpEF: heart failure with preserved ejection fraction; HFrEF: heart failure with reduced ejection fraction; SGLT2i: sodium-glucose cotransporter-2 inhibitor.

**Figure 3.**
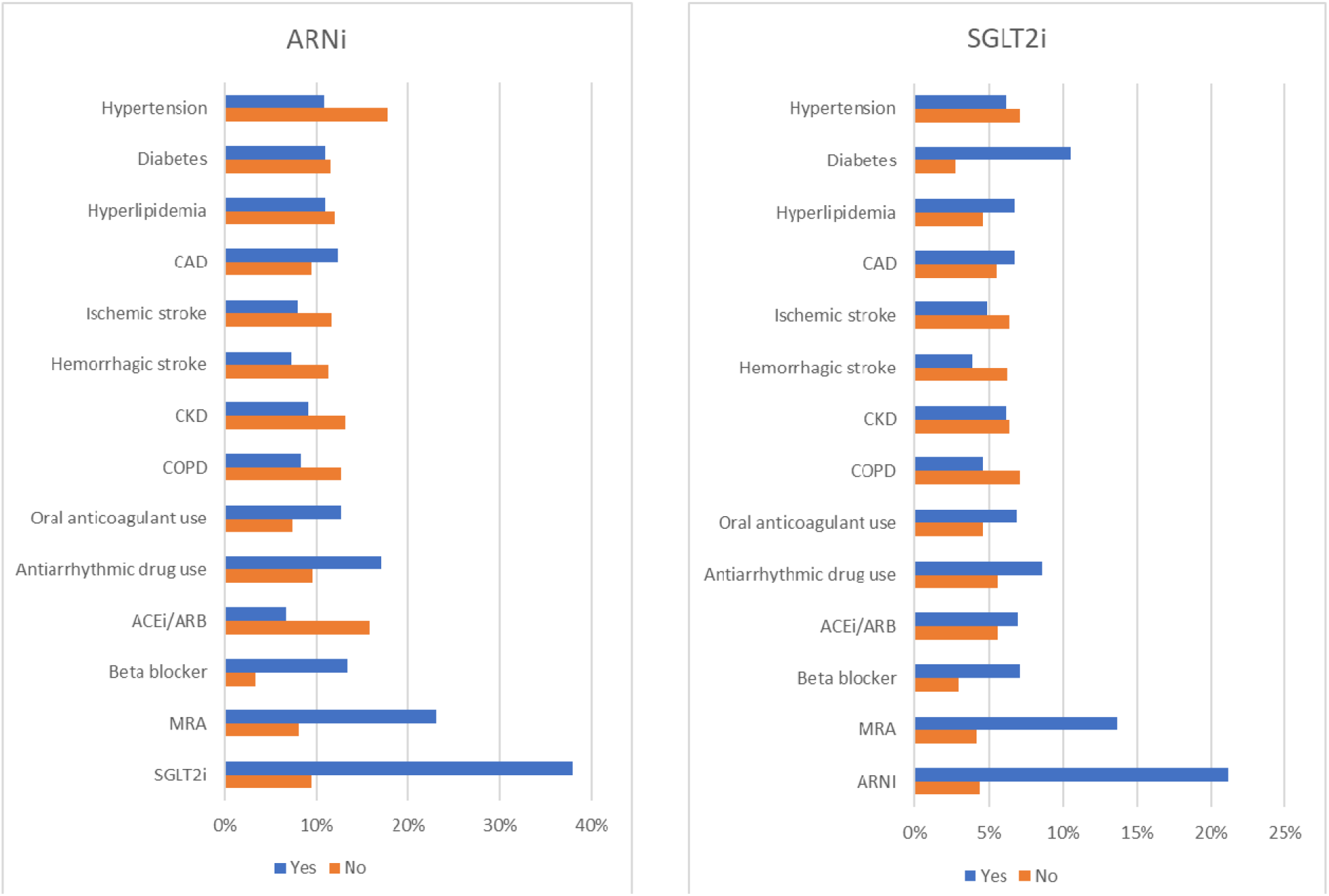
Prevalence of angiotensin receptor-neprilysin inhibitor and sodium-glucose cotransporter-2 inhibitor use among patients with atrial fibrillation and heart failure by presence of comorbidities and use of other relevant medications, MarketScan databases 2021-2022. ARNi: angiotensin receptor-neprilysin inhibitor; CAD: coronary artery disease; CKD: chronic kidney disease; COPD: chronic obstructive pulmonary disease; MRA: mineralocorticoid receptor antagonist; SGLT2i: sodium-glucose cotransporter-2 inhibitor.

## DISCUSSION

In a large contemporary sample of patients with AF and HF, we found that frequency of ARNi and SGLT2i use is low, even among patients with HFrEF, but increasing. We identified potential age and sex differences in the utilization of these medications, with female and older patients being less likely to fill prescriptions for ARNi or SGLT2i, in unadjusted analyses and also independently of clinical variables. Low use of SGLT2i in patients with HFpEF is likely influenced by this indication not being approved until early 2022.

Our results in the MarketScan databases are consistent with reports from other sources. A recent analysis of the Get With The Guidelines (GWTG)-HF registry reported that only 20% of patients hospitalized for HFrEF were prescribed an SGLT2i at discharge, with lower proportions among females and people older than 75 years of age.^14^ Important differences with our analysis include the inclusion of HFpEF in our sample, the restriction to patients with comorbid AF, and the use of information from filled outpatient prescriptions, a better marker of utilization than prescriptions at hospital discharge since not all HF patients fill their prescriptions after discharge.^19^ For example, in a different analysis of the GWTG-HF registry, more than one third of HFrEF patients discharged from the hospital with a new prescription for ARNi did not have any evidence of ARNi use in the 90-day period after discharge.^20^

The low frequency of ARNi and SGLT2i use in our study population underscore existing gaps in the use of guideline-directed medical therapy in patients with HF. In the CHAMP-HF registry, including 3,518 HFrEF patients enrolled between 2015 and 2017, less than 25% of patients were receiving triple therapy (ACEI/ARB/ARNI, beta-blocker, and MRA), and only 1% of eligible patients were receiving target doses of all three medications.^21^ Older age and comorbidities were associated with lower utilization. Given the impact of suboptimal HF medical therapy on negative outcomes, including HF admissions, the high prevalence of HF, and the known efficacy of guideline-recommended therapies, identifying and implementing interventions that improve use and adherence of these medications, including increasing affordability, should be a priority. How these interventions can be adapted to patients with comorbid HF and AF requires additional attention.

The present analysis has important strengths, including the large sample size, the focus on an understudied population, the use of real-world data, and the contemporary relevance of the data. These strengths should be tempered by relevant limitations, including the lack of information on race, ethnicity or socioeconomic status in the MarketScan databases and, therefore, the inability to evaluate the impact of these variables on ARNi and SGLT2i prescriptions. Furthermore, misclassification imposed by the use of healthcare claims to define the study population, predictors and endpoints, and the lack of objective information on ejection fraction and other HF-specific relevant variables to characterize the type and severity of disease can result in misclassification and biased results. Generalizability of these findings to patient populations not represented in MarketScan databases (e.g. Medicaid enrollees, uninsured) may be inadequate. Additionally, the unavailability of data after June 2022 limits our ability to properly evaluate use of SGLT2i in patients with HFpEF. Moreover, data reflects a specific time period in which the COVID-19 pandemic still had a major impact on healthcare delivery, which likely affected care of HF patients.

In conclusion, our analysis shows an important gap in the use of ARNi and SGLT2i in patients with comorbid HF and AF, identifies correlates of lower use, and reports on recent trends in prescriptions of these medications by HF type. This information could be used to develop targeted interventions to improve the management of patients with AF and HF. Future studies with more contemporary data should evaluate the intake of SGLT2i in patients with HFpEF, as well as the comparative effectiveness of these medications in real-world population.

## Supporting information

Supplemental Results

## Data Availability

Because of licensing restrictions, data and study materials cannot be made available to other investigators to reproduce results, but researchers may contact Merative to obtain and license the data.

---

**Supplemental Figure 1.**
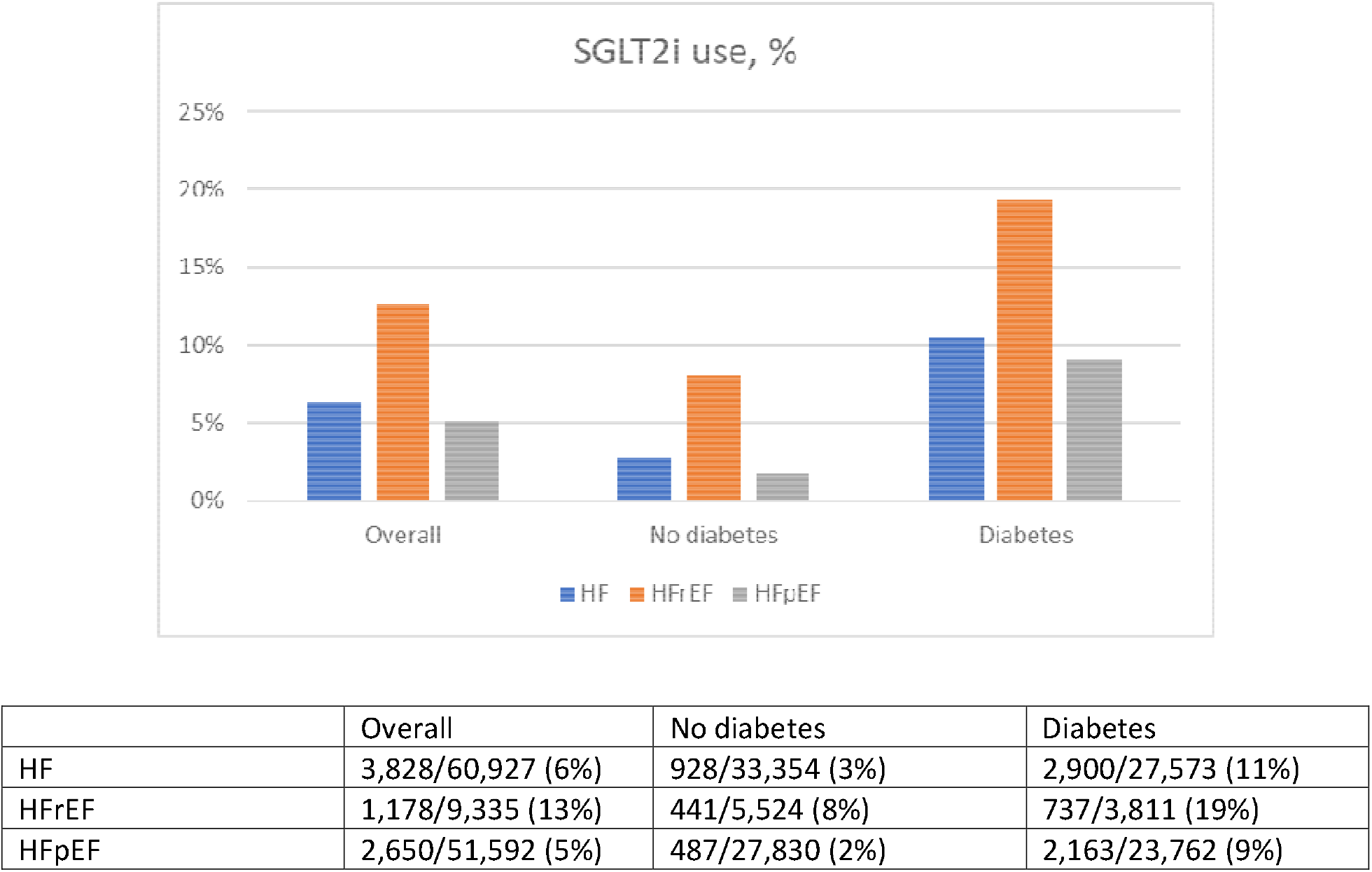
Prevalence of sodium-glucose cotransporter-2 inhibitors use among patients with atrial fibrillation and heart failure overall and by diabetes status, MarketScan databases 2021-2022. HFpEF: heart failure with preserved ejection fraction; HFrEF: heart failure with reduced ejection fraction SGLT2i: sodium-glucose cotransporter-2 inhibitor.

**Supplemental Figure 2.**
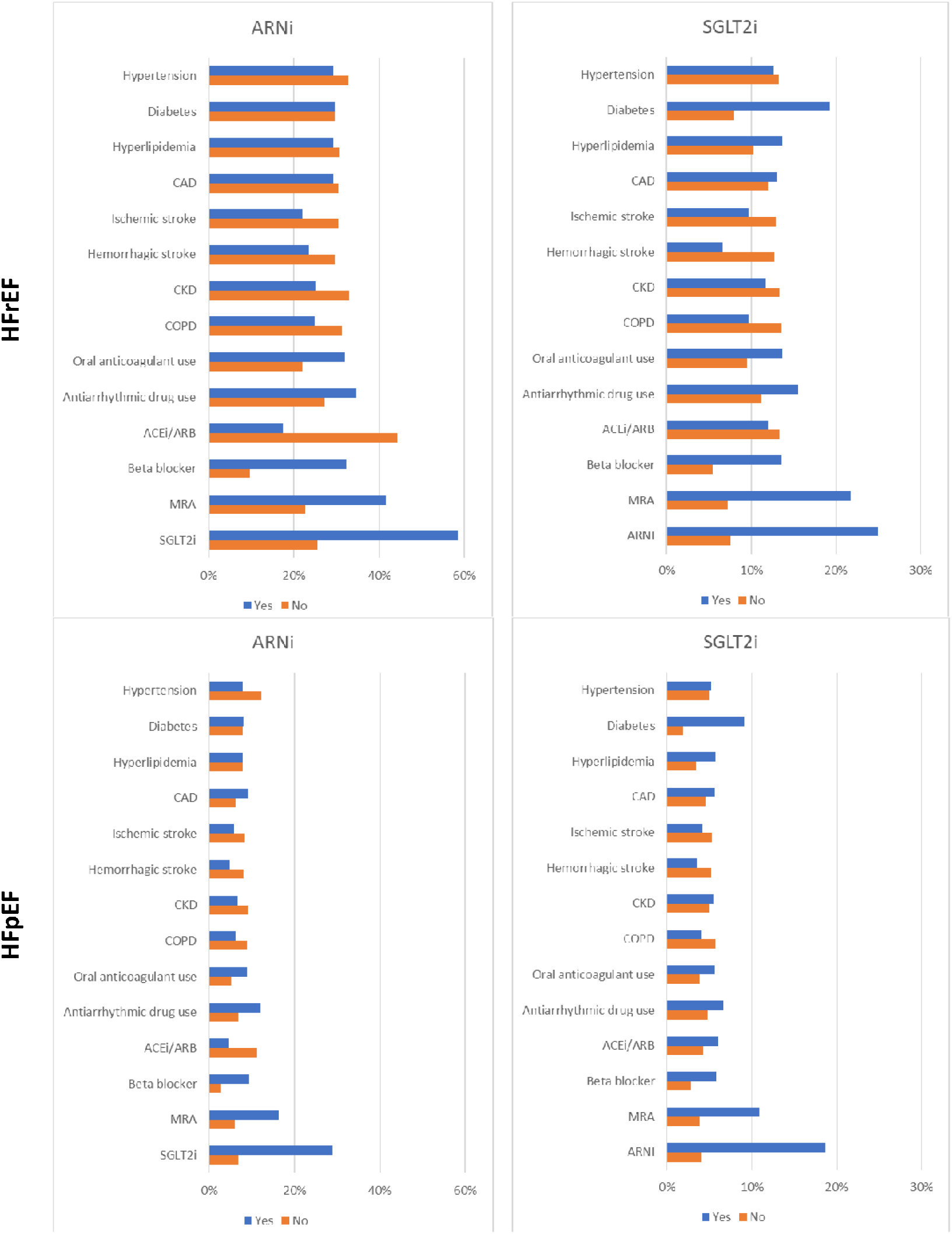
Prevalence of angiotensin receptor-neprilysin inhibitor and sodium-glucose cotransporter-2 inhibitor use among patients with atrial fibrillation and heart failure by presence of comorbidities and use of other relevant medications, by HF type, MarketScan databases 2021-2022. ARNi: angiotensin receptor-neprilysin inhibitor; CAD: coronary artery disease; CKD: chronic kidney disease; COPD: chronic obstructive pulmonary disease; MRA: mineralocorticoid receptor antagonist; SGLT2i: sodium-glucose cotransporter-2 inhibitor.

**Supplemental Table 1.**
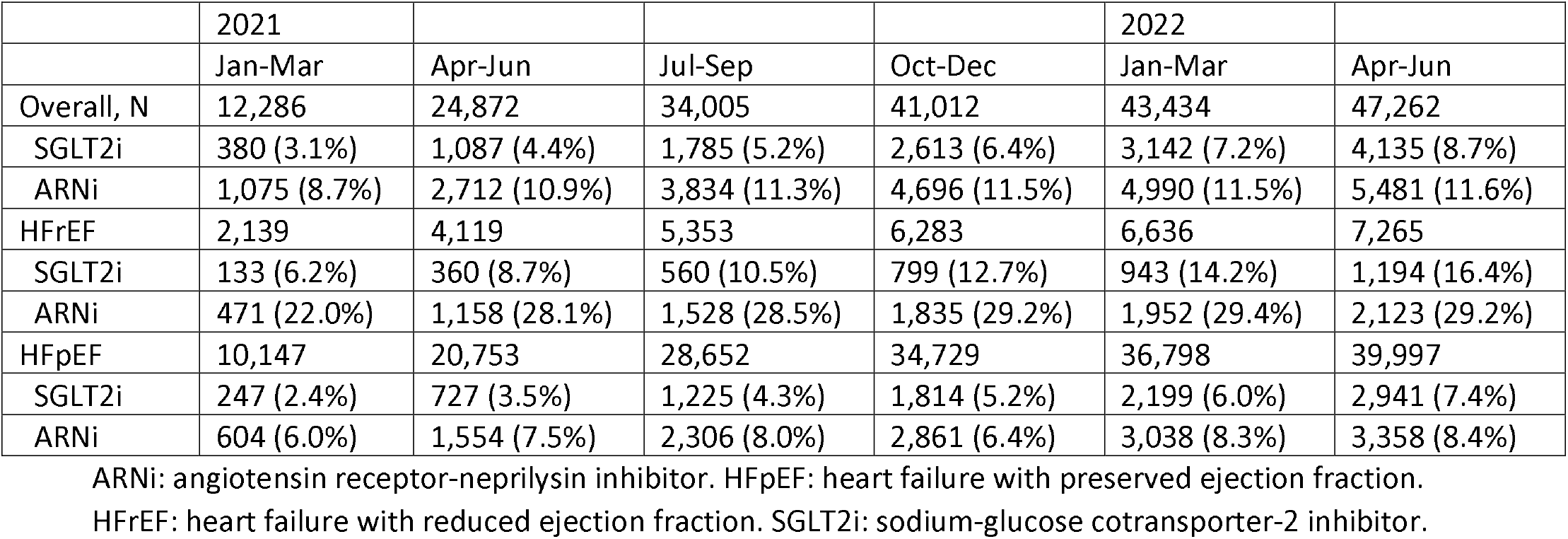
Prevalence of angiotensin receptor-neprilysin inhibitor and sodium-glucose cotransporter-2 inhibitor use among patients with AF and HF by quarter, MarketScan databases 2021-2022.

**Supplemental Table 2.**
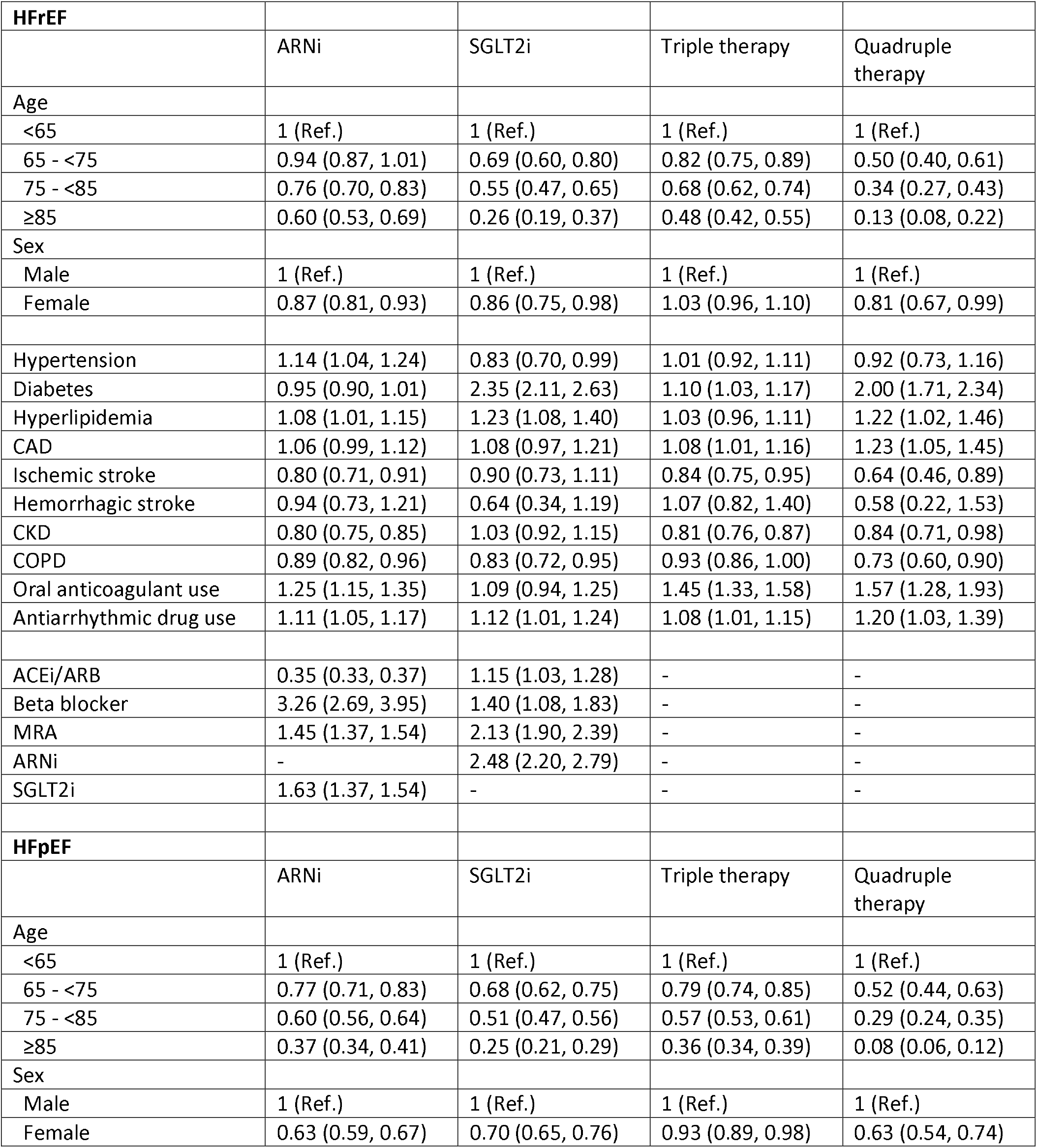
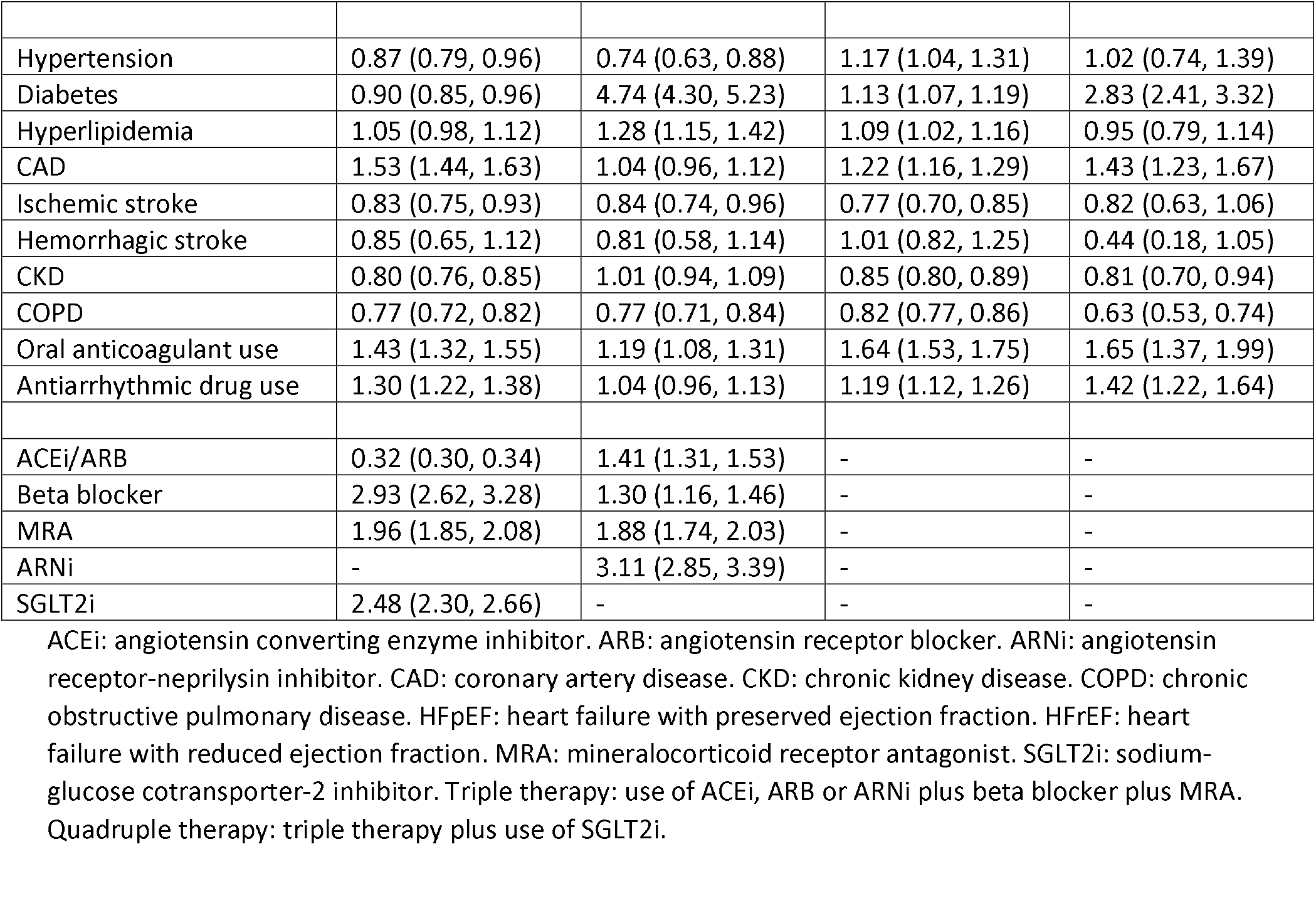
Predictors of use of angiotensin receptor-neprilysin inhibitor, sodium-glucose cotransporter-2 inhibitor, triple therapy and quadruple therapy in patients with AF and HF at time of co-diagnosis (+/-90 days) by HF type (HFrEF, HFpEF). Results correspond to relative risks and 95% confidence intervals from log-linear model (or Poisson regression when model not converging) including all variables in the table. MarketScan, 2021-2022.

**Supplemental Table 3.**
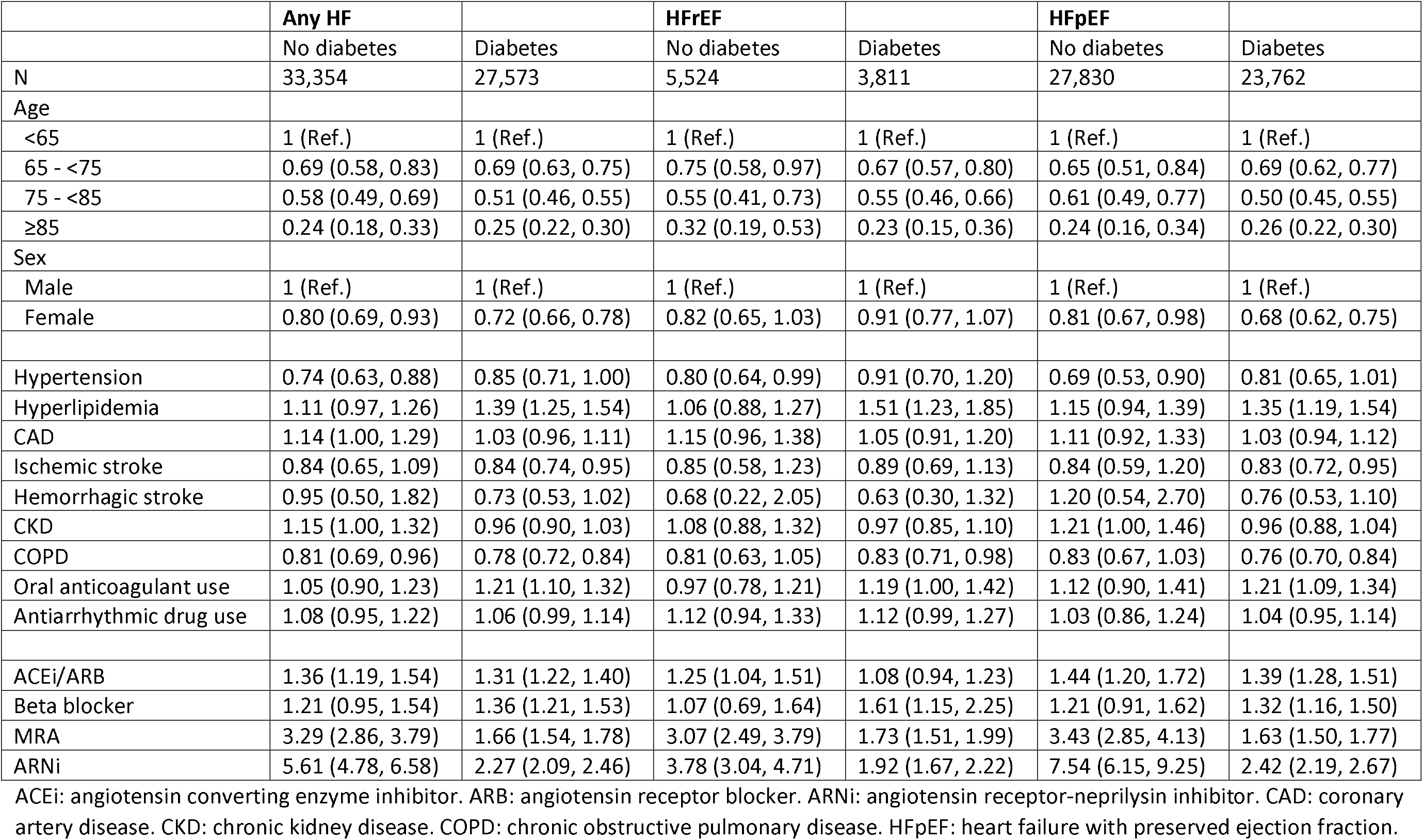
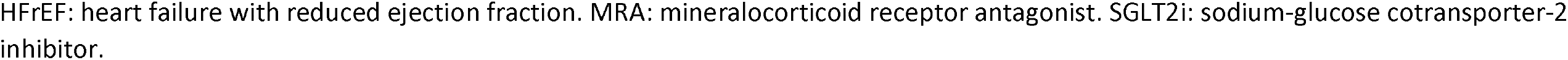
Predictors of use of sodium-glucose cotransporter-2 inhibitor in patients with AF and HF at time of co-diagnosis (+/-90 days) by diabetes status and HF type (HFrEF, HFpEF). Results correspond to relative risks and 95% confidence intervals from Poisson regression with robust variance estimation including all variables in the table. MarketScan, 2021-2022.

