## Supplemental Results for "Use of SGLT2i and ARNi in patients with atrial fibrillation and heart failure in 2021-2022: an analysis of real-world data"

Supplemental Figure 1. Prevalence of sodium-glucose cotransporter-2 inhibitors use among patients with atrial fibrillation and heart failure overall and by diabetes status, MarketScan databases 2021-2022. HFpEF: heart failure with preserved ejection fraction; HFrEF: heart failure with reduced ejection fraction; SGLT2i: sodium-glucose cotransporter-2 inhibitor.

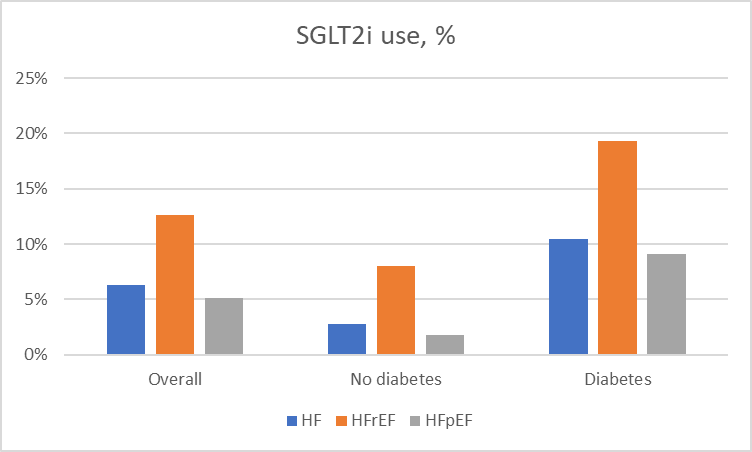

|  | Overall | No diabetes | Diabetes |
| --- | --- | --- | --- |
| HF | 3,828/60,927 (6%) | 928/33,354 (3%) | 2,900/27,573 (11%) |
| HFrEF | 1,178/9,335 (13%) | 441/5,524 (8%) | 737/3,811 (19%) |
| HFpEF | 2,650/51,592 (5%) | 487/27,830 (2%) | 2,163/23,762 (9%) |

Supplemental Figure 2. Prevalence of angiotensin receptor-neprilysin inhibitor and sodium-glucose cotransporter-2 inhibitor use among patients with atrial fibrillation and heart failure by presence of comorbidities and use of other relevant medications, by HF type, MarketScan databases 2021-2022. ARNi: angiotensin receptor-neprilysin inhibitor; CAD: coronary artery disease; CKD: chronic kidney disease; COPD: chronic obstructive pulmonary disease; MRA: mineralocorticoid receptor antagonist; SGLT2i: sodium-glucose cotransporter-2 inhibitor.

| **HFrEF** | 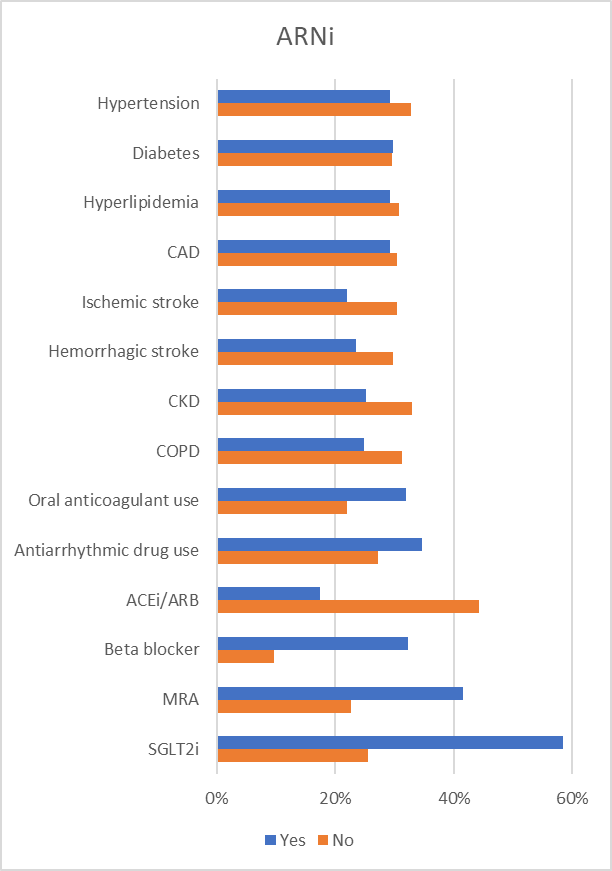 | 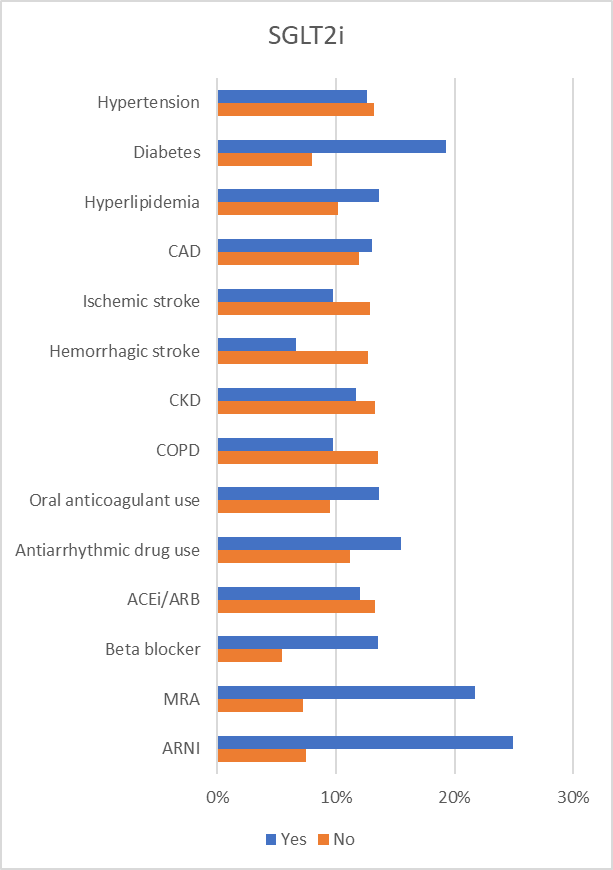 |
| --- | --- | --- |
| **HFpEF** | 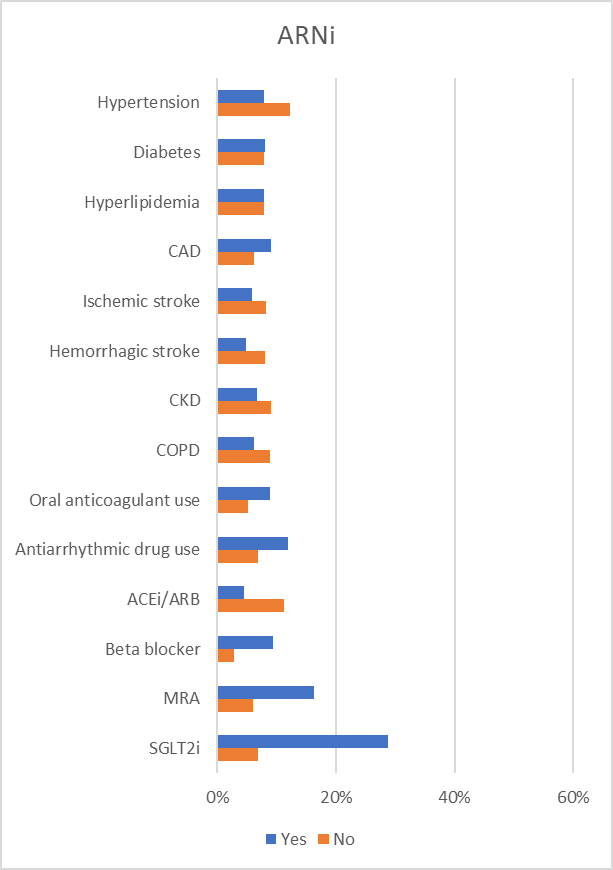 | 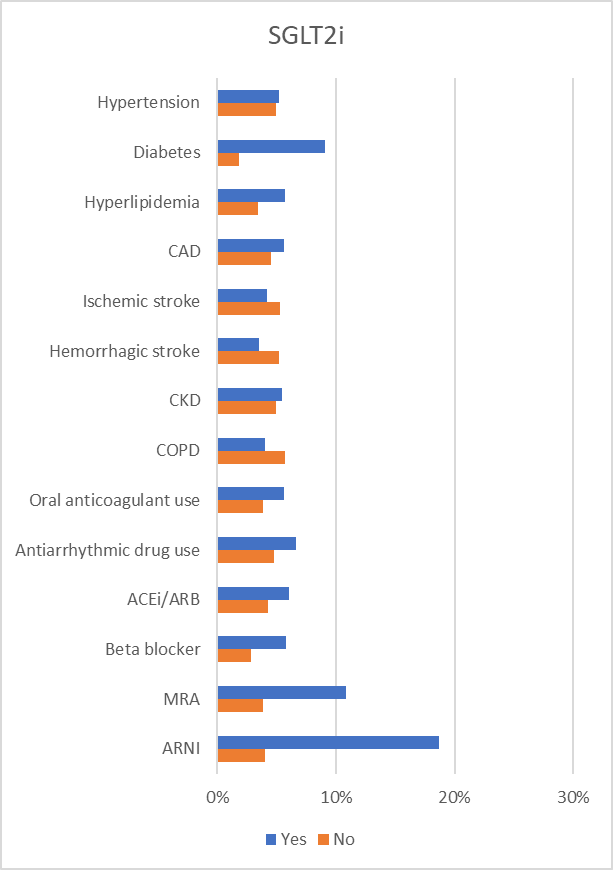 |

Supplemental Table 1. Prevalence of angiotensin receptor-neprilysin inhibitor and sodium-glucose cotransporter-2 inhibitor use among patients with AF and HF by quarter, MarketScan databases 2021-2022.

|  | 2021 |  |  |  | 2022 |  |
| --- | --- | --- | --- | --- | --- | --- |
|  | Jan-Mar | Apr-Jun | Jul-Sep | Oct-Dec | Jan-Mar | Apr-Jun |
| Overall, N | 12,286 | 24,872 | 34,005 | 41,012 | 43,434 | 47,262 |
| SGLT2i | 380 (3.1%) | 1,087 (4.4%) | 1,785 (5.2%) | 2,613 (6.4%) | 3,142 (7.2%) | 4,135 (8.7%) |
| ARNi | 1,075 (8.7%) | 2,712 (10.9%) | 3,834 (11.3%) | 4,696 (11.5%) | 4,990 (11.5%) | 5,481 (11.6%) |
| HFrEF | 2,139 | 4,119 | 5,353 | 6,283 | 6,636 | 7,265 |
| SGLT2i | 133 (6.2%) | 360 (8.7%) | 560 (10.5%) | 799 (12.7%) | 943 (14.2%) | 1,194 (16.4%) |
| ARNi | 471 (22.0%) | 1,158 (28.1%) | 1,528 (28.5%) | 1,835 (29.2%) | 1,952 (29.4%) | 2,123 (29.2%) |
| HFpEF | 10,147 | 20,753 | 28,652 | 34,729 | 36,798 | 39,997 |
| SGLT2i | 247 (2.4%) | 727 (3.5%) | 1,225 (4.3%) | 1,814 (5.2%) | 2,199 (6.0%) | 2,941 (7.4%) |
| ARNi | 604 (6.0%) | 1,554 (7.5%) | 2,306 (8.0%) | 2,861 (6.4%) | 3,038 (8.3%) | 3,358 (8.4%) |

ARNi: angiotensin receptor-neprilysin inhibitor. HFpEF: heart failure with preserved ejection fraction. HFrEF: heart failure with reduced ejection fraction. SGLT2i: sodium-glucose cotransporter-2 inhibitor.

Supplemental Table 2. Predictors of use of angiotensin receptor-neprilysin inhibitor, sodium-glucose cotransporter-2 inhibitor, triple therapy and quadruple therapy in patients with AF and HF at time of co-diagnosis (+/- 90 days) by HF type (HFrEF, HFpEF). Results correspond to relative risks and 95% confidence intervals from log-linear model (or Poisson regression when model not converging) including all variables in the table. MarketScan, 2021-2022.

| **HFrEF** |  |  |  |  |
| --- | --- | --- | --- | --- |
|  | ARNi | SGLT2i | Triple therapy | Quadruple therapy |
| Age |  |  |  |  |
| <65 | 1 (Ref.) | 1 (Ref.) | 1 (Ref.) | 1 (Ref.) |
| 65 - <75 | 0.94 (0.87, 1.01) | 0.69 (0.60, 0.80) | 0.82 (0.75, 0.89) | 0.50 (0.40, 0.61) |
| 75 - <85 | 0.76 (0.70, 0.83) | 0.55 (0.47, 0.65) | 0.68 (0.62, 0.74) | 0.34 (0.27, 0.43) |
| ≥85 | 0.60 (0.53, 0.69) | 0.26 (0.19, 0.37) | 0.48 (0.42, 0.55) | 0.13 (0.08, 0.22) |
| Sex |  |  |  |  |
| Male | 1 (Ref.) | 1 (Ref.) | 1 (Ref.) | 1 (Ref.) |
| Female | 0.87 (0.81, 0.93) | 0.86 (0.75, 0.98) | 1.03 (0.96, 1.10) | 0.81 (0.67, 0.99) |
| Hypertension | 1.14 (1.04, 1.24) | 0.83 (0.70, 0.99) | 1.01 (0.92, 1.11) | 0.92 (0.73, 1.16) |
| Diabetes | 0.95 (0.90, 1.01) | 2.35 (2.11, 2.63) | 1.10 (1.03, 1.17) | 2.00 (1.71, 2.34) |
| Hyperlipidemia | 1.08 (1.01, 1.15) | 1.23 (1.08, 1.40) | 1.03 (0.96, 1.11) | 1.22 (1.02, 1.46) |
| CAD | 1.06 (0.99, 1.12) | 1.08 (0.97, 1.21) | 1.08 (1.01, 1.16) | 1.23 (1.05, 1.45) |
| Ischemic stroke | 0.80 (0.71, 0.91) | 0.90 (0.73, 1.11) | 0.84 (0.75, 0.95) | 0.64 (0.46, 0.89) |
| Hemorrhagic stroke | 0.94 (0.73, 1.21) | 0.64 (0.34, 1.19) | 1.07 (0.82, 1.40) | 0.58 (0.22, 1.53) |
| CKD | 0.80 (0.75, 0.85) | 1.03 (0.92, 1.15) | 0.81 (0.76, 0.87) | 0.84 (0.71, 0.98) |
| COPD | 0.89 (0.82, 0.96) | 0.83 (0.72, 0.95) | 0.93 (0.86, 1.00) | 0.73 (0.60, 0.90) |
| Oral anticoagulant use | 1.25 (1.15, 1.35) | 1.09 (0.94, 1.25) | 1.45 (1.33, 1.58) | 1.57 (1.28, 1.93) |
| Antiarrhythmic drug use | 1.11 (1.05, 1.17) | 1.12 (1.01, 1.24) | 1.08 (1.01, 1.15) | 1.20 (1.03, 1.39) |
| ACEi/ARB | 0.35 (0.33, 0.37) | 1.15 (1.03, 1.28) | - | - |
| Beta blocker | 3.26 (2.69, 3.95) | 1.40 (1.08, 1.83) | - | - |
| MRA | 1.45 (1.37, 1.54) | 2.13 (1.90, 2.39) | - | - |
| ARNi | - | 2.48 (2.20, 2.79) | - | - |
| SGLT2i | 1.63 (1.37, 1.54) | - | - | - |
| **HFpEF** |  |  |  |  |
|  | ARNi | SGLT2i | Triple therapy | Quadruple therapy |
| Age |  |  |  |  |
| <65 | 1 (Ref.) | 1 (Ref.) | 1 (Ref.) | 1 (Ref.) |
| 65 - <75 | 0.77 (0.71, 0.83) | 0.68 (0.62, 0.75) | 0.79 (0.74, 0.85) | 0.52 (0.44, 0.63) |
| 75 - <85 | 0.60 (0.56, 0.64) | 0.51 (0.47, 0.56) | 0.57 (0.53, 0.61) | 0.29 (0.24, 0.35) |
| ≥85 | 0.37 (0.34, 0.41) | 0.25 (0.21, 0.29) | 0.36 (0.34, 0.39) | 0.08 (0.06, 0.12) |
| Sex |  |  |  |  |
| Male | 1 (Ref.) | 1 (Ref.) | 1 (Ref.) | 1 (Ref.) |
| Female | 0.63 (0.59, 0.67) | 0.70 (0.65, 0.76) | 0.93 (0.89, 0.98) | 0.63 (0.54, 0.74) |
| Hypertension | 0.87 (0.79, 0.96) | 0.74 (0.63, 0.88) | 1.17 (1.04, 1.31) | 1.02 (0.74, 1.39) |
| Diabetes | 0.90 (0.85, 0.96) | 4.74 (4.30, 5.23) | 1.13 (1.07, 1.19) | 2.83 (2.41, 3.32) |
| Hyperlipidemia | 1.05 (0.98, 1.12) | 1.28 (1.15, 1.42) | 1.09 (1.02, 1.16) | 0.95 (0.79, 1.14) |
| CAD | 1.53 (1.44, 1.63) | 1.04 (0.96, 1.12) | 1.22 (1.16, 1.29) | 1.43 (1.23, 1.67) |
| Ischemic stroke | 0.83 (0.75, 0.93) | 0.84 (0.74, 0.96) | 0.77 (0.70, 0.85) | 0.82 (0.63, 1.06) |
| Hemorrhagic stroke | 0.85 (0.65, 1.12) | 0.81 (0.58, 1.14) | 1.01 (0.82, 1.25) | 0.44 (0.18, 1.05) |
| CKD | 0.80 (0.76, 0.85) | 1.01 (0.94, 1.09) | 0.85 (0.80, 0.89) | 0.81 (0.70, 0.94) |
| COPD | 0.77 (0.72, 0.82) | 0.77 (0.71, 0.84) | 0.82 (0.77, 0.86) | 0.63 (0.53, 0.74) |
| Oral anticoagulant use | 1.43 (1.32, 1.55) | 1.19 (1.08, 1.31) | 1.64 (1.53, 1.75) | 1.65 (1.37, 1.99) |
| Antiarrhythmic drug use | 1.30 (1.22, 1.38) | 1.04 (0.96, 1.13) | 1.19 (1.12, 1.26) | 1.42 (1.22, 1.64) |
| ACEi/ARB | 0.32 (0.30, 0.34) | 1.41 (1.31, 1.53) | - | - |
| Beta blocker | 2.93 (2.62, 3.28) | 1.30 (1.16, 1.46) | - | - |
| MRA | 1.96 (1.85, 2.08) | 1.88 (1.74, 2.03) | - | - |
| ARNi | - | 3.11 (2.85, 3.39) | - | - |
| SGLT2i | 2.48 (2.30, 2.66) | - | - | - |

ACEi: angiotensin converting enzyme inhibitor. ARB: angiotensin receptor blocker. ARNi: angiotensin receptor-neprilysin inhibitor. CAD: coronary artery disease. CKD: chronic kidney disease. COPD: chronic obstructive pulmonary disease. HFpEF: heart failure with preserved ejection fraction. HFrEF: heart failure with reduced ejection fraction. MRA: mineralocorticoid receptor antagonist. SGLT2i: sodium-glucose cotransporter-2 inhibitor. Triple therapy: use of ACEi, ARB or ARNi plus beta blocker plus MRA. Quadruple therapy: triple therapy plus use of SGLT2i.

Supplemental Table 3. Predictors of use of sodium-glucose cotransporter-2 inhibitor in patients with AF and HF at time of co-diagnosis (+/- 90 days) by diabetes status and HF type (HFrEF, HFpEF). Results correspond to relative risks and 95% confidence intervals from Poisson regression with robust variance estimation including all variables in the table. MarketScan, 2021-2022.

|  | **Any HF** |  | **HFrEF** |  | **HFpEF** |  |
| --- | --- | --- | --- | --- | --- | --- |
|  | No diabetes | Diabetes | No diabetes | Diabetes | No diabetes | Diabetes |
| N | 33,354 | 27,573 | 5,524 | 3,811 | 27,830 | 23,762 |
| Age |  |  |  |  |  |  |
| <65 | 1 (Ref.) | 1 (Ref.) | 1 (Ref.) | 1 (Ref.) | 1 (Ref.) | 1 (Ref.) |
| 65 - <75 | 0.69 (0.58, 0.83) | 0.69 (0.63, 0.75) | 0.75 (0.58, 0.97) | 0.67 (0.57, 0.80) | 0.65 (0.51, 0.84) | 0.69 (0.62, 0.77) |
| 75 - <85 | 0.58 (0.49, 0.69) | 0.51 (0.46, 0.55) | 0.55 (0.41, 0.73) | 0.55 (0.46, 0.66) | 0.61 (0.49, 0.77) | 0.50 (0.45, 0.55) |
| ≥85 | 0.24 (0.18, 0.33) | 0.25 (0.22, 0.30) | 0.32 (0.19, 0.53) | 0.23 (0.15, 0.36) | 0.24 (0.16, 0.34) | 0.26 (0.22, 0.30) |
| Sex |  |  |  |  |  |  |
| Male | 1 (Ref.) | 1 (Ref.) | 1 (Ref.) | 1 (Ref.) | 1 (Ref.) | 1 (Ref.) |
| Female | 0.80 (0.69, 0.93) | 0.72 (0.66, 0.78) | 0.82 (0.65, 1.03) | 0.91 (0.77, 1.07) | 0.81 (0.67, 0.98) | 0.68 (0.62, 0.75) |
| Hypertension | 0.74 (0.63, 0.88) | 0.85 (0.71, 1.00) | 0.80 (0.64, 0.99) | 0.91 (0.70, 1.20) | 0.69 (0.53, 0.90) | 0.81 (0.65, 1.01) |
| Hyperlipidemia | 1.11 (0.97, 1.26) | 1.39 (1.25, 1.54) | 1.06 (0.88, 1.27) | 1.51 (1.23, 1.85) | 1.15 (0.94, 1.39) | 1.35 (1.19, 1.54) |
| CAD | 1.14 (1.00, 1.29) | 1.03 (0.96, 1.11) | 1.15 (0.96, 1.38) | 1.05 (0.91, 1.20) | 1.11 (0.92, 1.33) | 1.03 (0.94, 1.12) |
| Ischemic stroke | 0.84 (0.65, 1.09) | 0.84 (0.74, 0.95) | 0.85 (0.58, 1.23) | 0.89 (0.69, 1.13) | 0.84 (0.59, 1.20) | 0.83 (0.72, 0.95) |
| Hemorrhagic stroke | 0.95 (0.50, 1.82) | 0.73 (0.53, 1.02) | 0.68 (0.22, 2.05) | 0.63 (0.30, 1.32) | 1.20 (0.54, 2.70) | 0.76 (0.53, 1.10) |
| CKD | 1.15 (1.00, 1.32) | 0.96 (0.90, 1.03) | 1.08 (0.88, 1.32) | 0.97 (0.85, 1.10) | 1.21 (1.00, 1.46) | 0.96 (0.88, 1.04) |
| COPD | 0.81 (0.69, 0.96) | 0.78 (0.72, 0.84) | 0.81 (0.63, 1.05) | 0.83 (0.71, 0.98) | 0.83 (0.67, 1.03) | 0.76 (0.70, 0.84) |
| Oral anticoagulant use | 1.05 (0.90, 1.23) | 1.21 (1.10, 1.32) | 0.97 (0.78, 1.21) | 1.19 (1.00, 1.42) | 1.12 (0.90, 1.41) | 1.21 (1.09, 1.34) |
| Antiarrhythmic drug use | 1.08 (0.95, 1.22) | 1.06 (0.99, 1.14) | 1.12 (0.94, 1.33) | 1.12 (0.99, 1.27) | 1.03 (0.86, 1.24) | 1.04 (0.95, 1.14) |
| ACEi/ARB | 1.36 (1.19, 1.54) | 1.31 (1.22, 1.40) | 1.25 (1.04, 1.51) | 1.08 (0.94, 1.23) | 1.44 (1.20, 1.72) | 1.39 (1.28, 1.51) |
| Beta blocker | 1.21 (0.95, 1.54) | 1.36 (1.21, 1.53) | 1.07 (0.69, 1.64) | 1.61 (1.15, 2.25) | 1.21 (0.91, 1.62) | 1.32 (1.16, 1.50) |
| MRA | 3.29 (2.86, 3.79) | 1.66 (1.54, 1.78) | 3.07 (2.49, 3.79) | 1.73 (1.51, 1.99) | 3.43 (2.85, 4.13) | 1.63 (1.50, 1.77) |
| ARNi | 5.61 (4.78, 6.58) | 2.27 (2.09, 2.46) | 3.78 (3.04, 4.71) | 1.92 (1.67, 2.22) | 7.54 (6.15, 9.25) | 2.42 (2.19, 2.67) |

ACEi: angiotensin converting enzyme inhibitor. ARB: angiotensin receptor blocker. ARNi: angiotensin receptor-neprilysin inhibitor. CAD: coronary artery disease. CKD: chronic kidney disease. COPD: chronic obstructive pulmonary disease. HFpEF: heart failure with preserved ejection fraction. HFrEF: heart failure with reduced ejection fraction. MRA: mineralocorticoid receptor antagonist. SGLT2i: sodium-glucose cotransporter-2 inhibitor.
